# Variance quantitative trait loci reveal gene-gene interactions which alter blood traits

**DOI:** 10.1101/2024.09.18.24313883

**Authors:** Yash Pershad, Hannah Poisner, Robert W Corty, Jacklyn N Hellwege, Alexander G Bick

## Abstract

Gene-gene (GxG) interactions play an important role in human genetics, potentially explaining part of the “missing heritability” of polygenic traits and the variable expressivity of monogenic traits. Many GxG interactions have been identified in model organisms through experimental breeding studies, but they have been difficult to identify in human populations. To address this challenge, we applied two complementary variance QTL (vQTL)-based approaches to identify GxG interactions that contribute to human blood traits and blood-related disease risk. First, we used the previously validated genome-wide scale test for each trait in ∼450,000 people in the UK Biobank and identified 4 vQTLs. Genome-wide GxG interaction testing of these vQTLs enabled discovery of novel interactions between (1) *CCL24* and *CCL26* for eosinophil count and plasma CCL24 and CCL26 protein levels and (2) *HLA-DQA1* and *HLA-DQB1* for lymphocyte count and risk of celiac disease, both of which replicated in ∼140,000 NIH All of Us and ∼70,000 Vanderbilt BioVU participants. Second, we used a biologically informed approach to search for vQTL in disease-relevant genes. This approach identified (1) a known interaction for hemoglobin between two pathogenic variants in *HFE* which cause hereditary hemochromatosis and alters risk of cirrhosis and (2) a novel interaction between the *JAK2* 46/1 haplotype and a variant on chromosome 14 which modifies platelet count, *JAK2* V617F clonal hematopoiesis, and risk of polycythemia vera. This work identifies novel disease-relevant GxG interactions and demonstrates the utility of vQTL-based approaches in identifying GxG interactions relevant to human health at scale.

## Main

In gene-gene interactions, or epistasis, the effect of a variant depends on the genotype at another locus.^1^ Gene-gene interactions are significant in human genetics, as they may explain missing heritability of polygenic traits and variable expressivity of monogenic diseases.^2–4^ Systematically studying gene-gene interactions in humans is challenging due to the lack of sufficient power and the curse of multiplicity.^5^ Therefore, prior work has aimed to prioritize which variants to test for interactions.^6–11^ These studies have identified that genetic variants associated with variance of a trait – variance quantitative trait loci (vQTLs) – are more likely to be involved in a gene-gene or gene-environment interactions. Here, by prioritizing vQTLs – first genome-wide and second in disease-relevant genes – we identify gene-gene interactions altering blood traits (hemoglobin, platelet count, lymphocyte count, and eosinophil count) and disease risk among up to 462,468 people in the UK Biobank (**Supplementary Fig 1**).

We began by evaluating whether a vQTL approach can identify a known gene-gene interaction between *PNPLA3* p.I148M (rs738409) and rs72613567:TA in *HSD17B13* as a proof of concept.^12^ We modeled the variance effects of 76 SNPs with minor allele frequency (mAF) > 10% in *PNPLA3* for aspartate aminotransferase using double generalized linear models (dglm).^13^ The p.I148M variant was a vQTL, with the most significant dispersion of all extracted variants in *PNPLA3* (β_dispersion_ = 0.18, 95% confidence interval (CI) = [0.14, 0.22], P = 6.74×10^-18^) (**Supplementary Fig 2**).

In order to find genome-wide vQTLs for hemoglobin, platelet count, lymphocyte count, and eosinophil count, we performed the genome-wide Scale test^7^ for each trait in the UK Biobank. A variant was identified as a vQTL if it had a genome-wide significant association with the square of the residual of a trait without a significant mean effect after multiple-hypothesis correction (See **Methods**). We identified four vQTLs, with two for lymphocyte count and two for eosinophil count (**Supplementary Table 1**). For each of these four variants, we performed a genome-wide association study for gene-gene interactions with the trait of interest using REGENIE^14^ and found two significant interactions.

The first gene-gene interaction discovered was between vQTL rs10281069 and rs112610805 for eosinophil count (**Fig 1a**, β_int_ = 0.0025 10^9^ cells/L, 95% CI = [0.0013, 0.0037], P = 5.44×10^-5^). rs10281069 is a known protein expression quantitative trait locus of *CCL24*, while rs112610805 is a known expression quantitative trait locus (eQTL) of CCL26 levels.^15^ CCL24 and CCL26 are both chemokines expressed in blood that affect eosinophil recruitment and activation.^16^ This positive interaction is suppressing, as in the complete model with the interaction term, eosinophil count is negatively associated with both rs10281069 (β = -0.0035 10^9^ cells/L, 95% CI = [- 0.0046, -0.0025], P = 4.65×10^-11^) and rs112610805 (β = -0.0021 10^9^ cells/L, 95% CI = [-0.0028, -0.0013], P = 8.29×10^-8^). The variants are in linkage equilibrium (**Fig 1b**; D’ = 0.28, R^2^ = 0.04). We observed the same positive suppressing interaction among 50,092 individuals in BioVU (β_int_ = 0.006 10^9^ cells/L, 95% CI = [0.002, 0.008], P = 0.001) and 138,860 individuals in All of Us (β_int_ = 0.0029 10^9^ cells/L, 95% CI = [0.0013, 0.0049], P = 2.42×10^-6^).

**Fig. 1:**
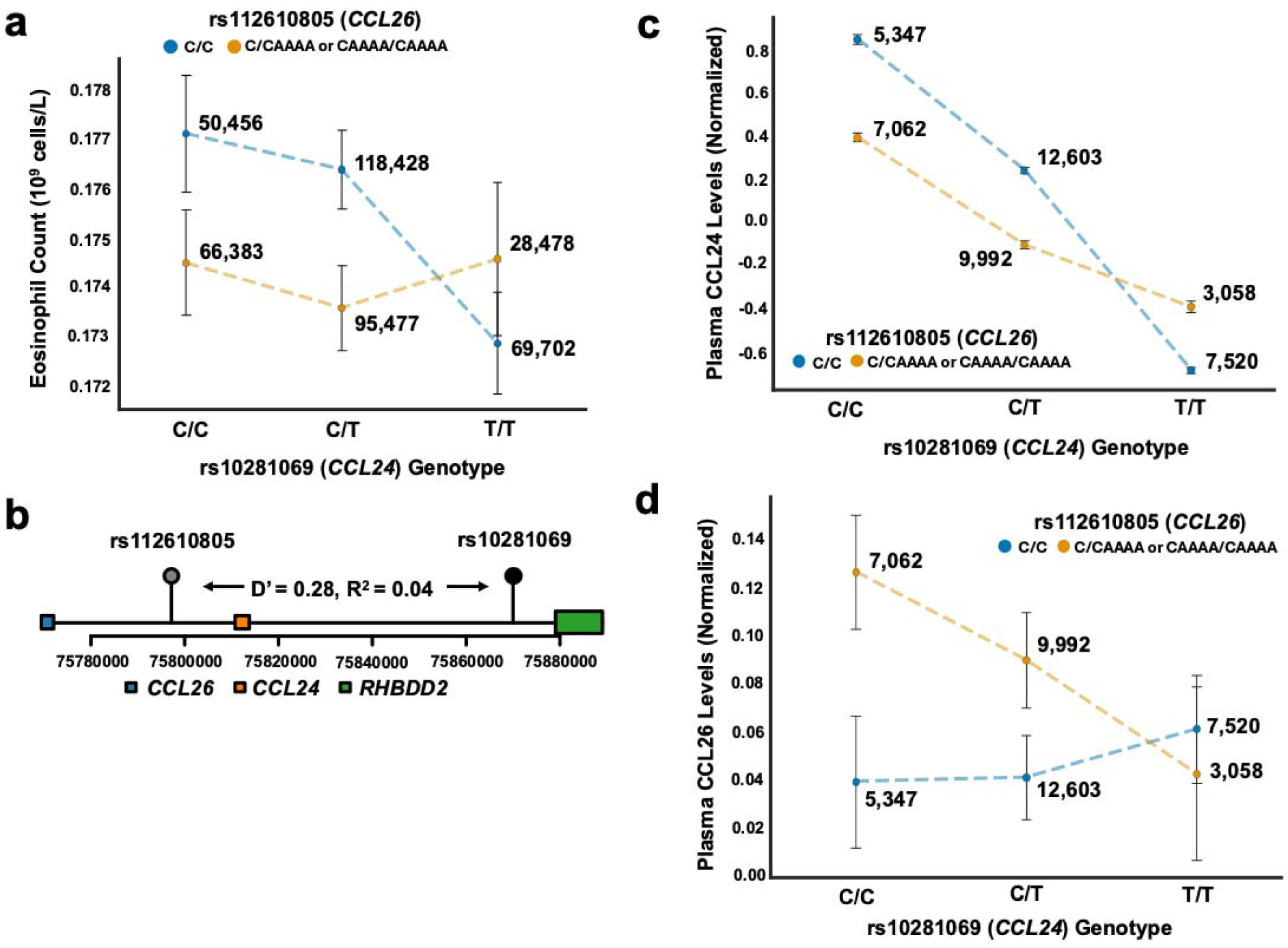
Genetic variants in CCL24 and CCL26 affect eosinophil count and plasma chemokine levels. a) Eosinophil count (10^9^ cells/L) stratified by genotypes at rs10281069 (*CCL24*) and rs112610805 (*CCL26*). The x-axis shows rs10281069 genotypes (C/C, C/T, T/T), with separate lines for rs112610805 genotypes (C/C in blue, C/CAAAA or CAAAA/CAAAA in orange). The y-axis shows mean eosinophil count. Error bars represent 95% confidence intervals (1.96*SE). Numbers beside each point indicate sample sizes for each genotype combination. b) Genomic context of rs10281069 and rs112610805. The plot shows the relative positions of *CCL24*, *CCL26*, and *RHBDD2* genes. The two variants are in linkage disequilibrium (D’ = 0.28, R² = 0.04). The x-axis shows the base position on chromosome 7. c) Plasma CCL24 levels (normalized) stratified by rs10281069 (*CCL24*) and rs112610805 (*CCL26*) genotypes. The x-axis shows rs10281069 genotypes, with colors representing rs112610805 genotypes as in (a). Sample sizes are indicated beside each box. Error bars represent 95% confidence intervals (1.96*SE). d) Plasma CCL26 levels (normalized) stratified by rs10281069 (*CCL24*) and rs112610805 (*CCL26*) genotypes similar to (c).

To understand the mechanism of the interaction, we tested for an interaction between the variants in *CCL24* and *CCL26* and chemokine abundance using plasma proteomics data from 50,562 participants in the UK Biobank.^17^ Both rs112610805 and rs10281069 were associated with lower plasma CCL24 (**Supplementary Table 2**) and positively interact (**Fig 1c**, β_int_ = 0.37, 95% CI = [0.35, 0.39], P = 4.99×10^-247^). For CCL26, rs112610805 and rs10281069 were associated with higher plasma CCL26 (**Supplementary Table 2**), and we observed a compensatory negative interaction (**Fig 1d**, β_int_ = -0.05, 95% CI = [-0.08, -0.02], P = 2.89×10^-4^). These data suggest that the suppressive interaction between rs10281069 and rs112610805 on eosinophil count arise from their opposing effects on CCL24 and CCL26 chemokine levels.

The second gene-gene interaction from the Scale test was between vQTL rs3819720 and rs4713570, affecting lymphocyte count. The variants rs3819720 and rs4713570 are known to associate with both splicing and expression for *HLA-DQB1*^18^ and *HLA-DQA1*^19^, respectively. Both rs3819720 and rs4713570 have a positive association with lymphocyte count in UK Biobank (**Supplementary Table 3**). Their interaction is significant and negative (**Fig 2a**, β_int_ = - 0.03 10^9^ cells/L, 95% CI = [-0.04, -0.02], P = 5.85×10^-15^). These variants are both on chromosome 6 and are in linkage equilibrium (**Fig 2b**, D’ = 0.0608, R^2^ = 0.0003). We observed the same relationship – a significant negative interaction for lymphocyte count – in 52,823 people in BioVU (β_int_ = -0.02 10^9^ cells/L, 95% CI = [-0.03, -0.0067], P = 0.003) and in 138,912 people in All of Us (β_int_ = -0.05 10^9^ cells/L, 95% CI = [-0.007, -0.0009], P = 0.02).

**Fig 2:**
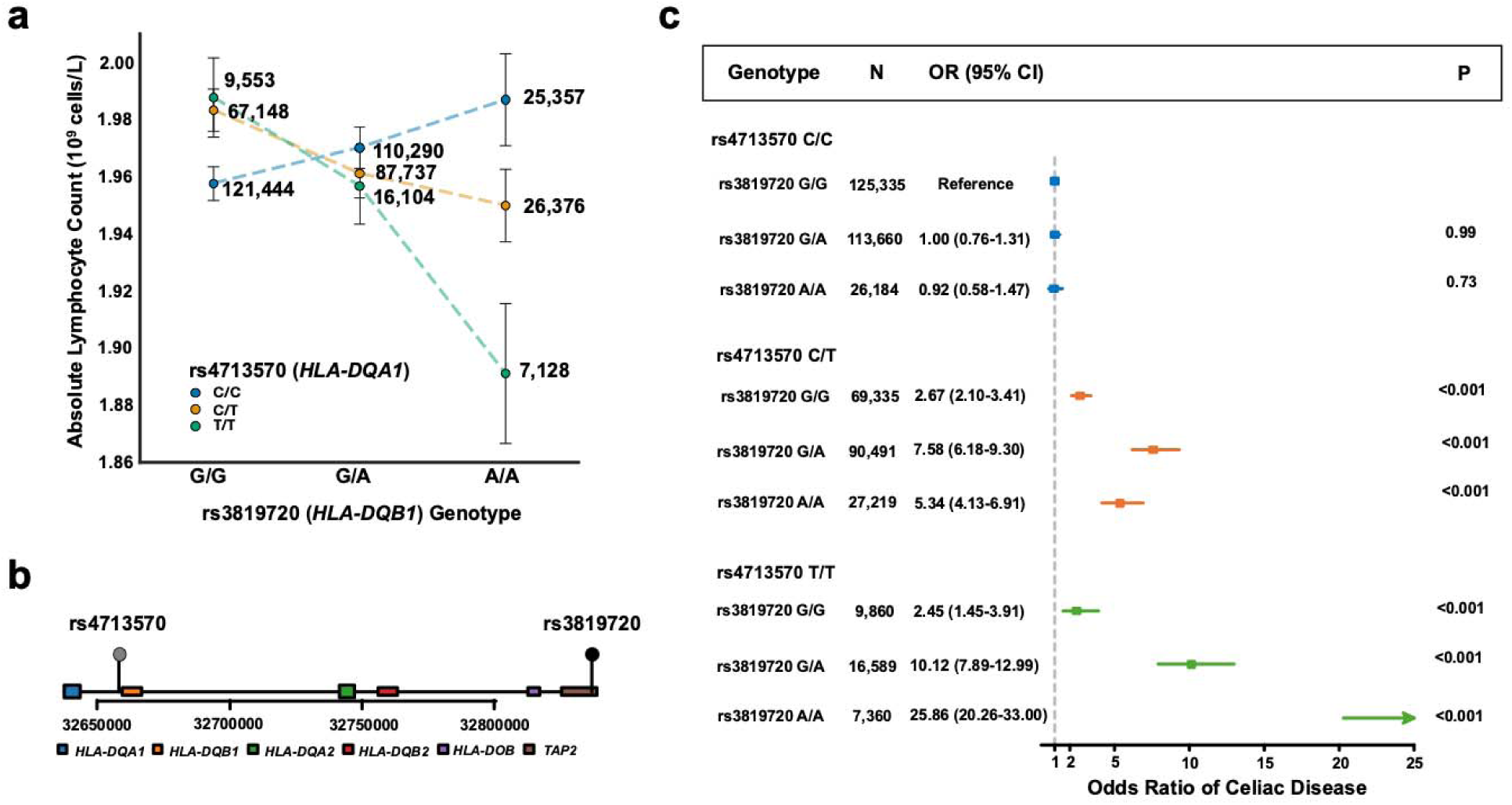
HLA-DQA1 and HLA-DQB1 variants affect absolute lymphocyte count and celiac disease risk. a) Absolute lymphocyte count (10^9^ cells/L) stratified by genotypes at rs3819720 (an expression quantitative trait locus of *HLA-DQB1*) and rs4713570 (an expression quantitative trait locus of *HLA-DQA1*). The x-axis shows rs3819720 genotypes (G/G, G/A, A/A), with separate lines for rs4713570 genotypes (C/C in blue, C/T in orange, T/T in green). The y-axis shows mean absolute lymphocyte count (10^9^ cells/L). Error bars represents 95% confidence intervals (1.96*SE). Numbers beside each point indicate sample sizes for each genotype combination. b) Genomic context of rs4713570 and rs3819720. The plot shows the relative positions of *HLA-DQA1*, *HLA-DQB1,* and neighboring HLA genes (*TAP2*, *HLA-DOB*, *HLA-DQB2*, *HLA-DQA2*) on chromosome 6. The x-axis shows the base position. c) Forest plot of odds ratios for celiac disease risk by genotype combinations of rs3819720 and rs4713570. The x-axis shows the odds ratio. For each genotype combination, the plot shows the number of individuals (N), odds ratio with 95% confidence interval (95% CI), and P-value (P). The rs3819720 G/G and rs4713570 C/C combination serves as the reference group.

*HLA-DQA1* and *HLA-DQB1* are genes that encode the alpha and beta chains of the HLA-DQ heterodimer, a key component of the major histocompatibility class II complex. Different combinations of *HLA-DQA1* and *HLA-DQB1* alleles can lead to preferential binding of specific peptides. Both variants are associated with autoimmune diseases such as type 1 diabetes and celiac disease.^20,21^ The suppressing interaction between rs3819720 and rs4713570 on lymphocyte count may be a result of a negative feedback mechanism in lymphocyte proliferation or formation of suboptimal HLA-DQ heterodimers when both variants are present.

As both rs3819720 and rs4713570 are associated with increased risk of celiac disease, we then sought to determine in the UK Biobank whether the variants interact to alter risk of celiac disease. rs3819720 and rs4713570 were both positively associated with prevalent celiac disease. The interaction between rs3819720 and rs4713570 for prevalent celiac disease was significant and enhancing (β_int_= 0.31, 95% CI = [0.21, 0.42], P = 1.70×10^-5^). Among homozygotes for the risk allele for rs4713570, the odds of celiac disease increased from 2.45 (95% CI = [1.45, 3.91], P = 1.69×10^-4^) for homozygotes for the rs3819720 reference allele to 25.86 (95% CI = [20.26, 33.00], P = 1.65×10^-150^) for homozygotes for the rs3819720 alternate allele (**Fig 2c**). These data together suggest that variants in *HLA-DQA1* and *HLA-DQB1* interact to reduce lymphocyte count in the blood and increase celiac disease risk possibly due to effects on the HLA-DQ heterodimer.

The scale test identified two other variants – rs340809 and rs13395354– as vQTLs for eosinophils count and lymphocyte count respectively (**Supplementary Table 1**). rs340809 is an intronic variant in *IL5RA*, which encodes a subunit of the IL-5 receptor; the IL-5 receptor binds IL-5, which is the major activator of eosinophils. rs13395354 overlaps with a promoter region for *BCL2L11* in five T-cell types and fetal thymus cells.^19^ *BCL2L11* encodes BIM (Bcl-2 Interacting Mediator of cell death), which is a pro-apoptotic protein expressed in lymphocytes.

Despite the variants’ biologic plausibility, we found no significant gene-gene interactions for these vQTLs. We also found no gene-environment interactions using previously defined 506 exposures by Hillary et al.^11^ These two vQTLs may interact with an unmeasured environmental factor or rare genetic variants, or they may represent false positives due to their suggestive mean effects (**Supplementary Table 1**).

While the Scale test allows us to find vQTLs in an unbiased manner genome-wide, the burden of multiple-hypothesis correction may limit our ability to find vQTLs. Therefore, we used a biologically informed approach to search for vQTL in genes relevant to monogenic diseases which affect blood traits (**Supplementary Table 4**). We only evaluated variants with a minor allele frequency (mAF) > 10% within these select genes to ensure sufficient power. We found significant vQTLs after Bonferroni correction for *JAK2*, *RUNX1*, *IL33*, and *HFE*, but not for *CHEK2* or *ENG* (**Supplementary Table 4)**. For these vQTLs, we performed a genome-wide test in the UK Biobank using the trait of interest for gene-gene interactions as described above (**Supplementary Table 4)**.^14^

First, we investigated the interaction hits for *HFE* and hemoglobin. Each of the vQTLs in *HFE* for hemoglobin interacted with one variant, rs4645. rs4645 had an enhancing interaction on the positive effect of the vQTL rs2858993 (**Fig 3a**, β_int_= 0.067 g/dL, 95% CI = [0.060, 0.075], P = 1.09 × 10^-71^). Moreover, rs4645 had a significant positive interaction with the two other vQTLs within *HFE* (rs12346 and rs2794719) for hemoglobin (**Supplementary Table 6**). Given that deleterious mutations in *HFE* cause type 1 hereditary hemochromatosis, we then tested whether the interaction also applied to the two most common pathogenic variants in *HFE*: C282Y (rs1800562) and H63D (rs1799945).^22^ We modeled the interaction between rs4645 and pathogenic allele count (i.e., compound heterozygotes carry two pathogenic alleles). The interaction between rs4645 and pathogenic allele count was significant (**Fig 3b**, β_int_ = 0.044 g/dL, 95% CI = [0.037, 0.053], P = 3.24×10^-27^), as was the interaction between rs4645 and each pathogenic variant separately (H63D: β_int_ = 0.017 g/dL, 95% CI = [0.0036, 0.030], P = 0.01; C282Y: β_int_ = 0.023 g/dL, 95% CI = [0.0066, 0.038], P = 5.45 x10^-3^). We observed the same positive interaction between rs4645 and pathogenic variant count among 70,623 individuals in BioVU (β_int_ = 0.075 10^9^ cells/L, 95% CI = [0.037, 0.112], P =1.41×10^-4^) and 138,940 individuals in All of Us (β_int_ = 0.11 10^9^ cells/L, 95% CI = [0.05, 0.17], P =2.29×10^-4^).

**Fig. 3:**
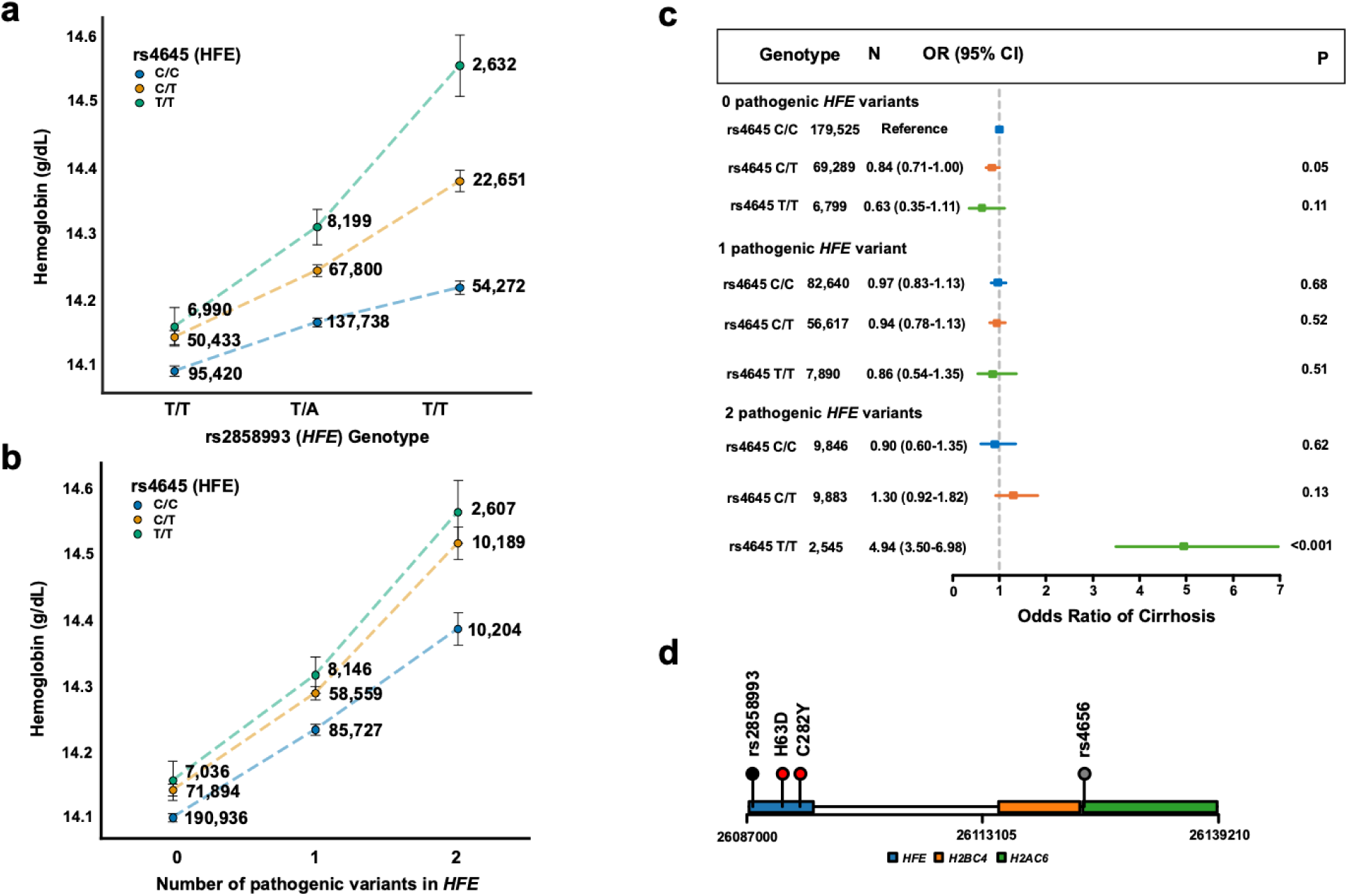
HFE variants affect hemoglobin levels and cirrhosis risk. a) Hemoglobin levels (g/dL) stratified by genotypes at rs2858993 and rs4645 in *HFE*. The x-axis shows rs2858993 genotypes (T/T, T/A, A/A), with separate lines for rs4645 genotypes (C/C in blue, C/T in orange, T/T in green). The y-axis shows mean hemoglobin levels. Error bars represent 95% confidence intervals (1.96*SE). Numbers beside each point indicate sample sizes for each genotype combination. b) Hemoglobin levels (g/dL) stratified by the number of pathogenic variants in *HFE* (0, 1, or 2). The x-axis shows the number of pathogenic variants, with separate lines for rs4645 genotypes (C/C in blue, C/T in orange, T/T in green). The y-axis shows mean hemoglobin levels. Error bars represent 95% confidence intervals (1.96*SE). Numbers beside each point indicate sample sizes for each category. c) Forest plot of odds ratios for cirrhosis risk by number of pathogenic *HFE* variants and rs4645 genotype. The x-axis shows the odds ratio of cirrhosis. For each combination, the plot shows the number of individuals (N), odds ratio with 95% confidence interval (CI), and P-value (P). The group with 0 pathogenic *HFE* variants serves as the reference. d) Genomic context of rs4656 and rs2858993 in *HFE*. The plot shows the relative positions of these variants within the *HFE* gene, including the locations of the H63D and C282Y mutations. Neighboring genes (*H2BC4*, *H2AC6*) are also shown. The x-axis indicates the base position on the chromosome.

We then compared the prevalence of liver cirrhosis – a complication of iron overload in hemochromatosis – stratified by alternate allele copies for rs4645 and pathogenic alleles in the UK Biobank. While rs4645 allele count was not associated with prevalent liver cirrhosis (β = 0.0083, 95% CI = [-0.094, 0.11], P = 0.87), the interaction of rs4645 and pathogenic allele count enhanced risk of prevalent liver cirrhosis (β_int_ = 0.48, 95% CI = [0.33, 0.62], P < 0.001). Among individuals with two pathogenic alleles, the odds of liver cirrhosis was 4.94 (95% CI: [3.50, 6.98], P < 0.001) for those with two alternate alleles for rs4645. Odds decreased to 1.30 (95% CI: [0.92,1.82], P = 0.13) for individuals with one alternate rs4645 allele and 0.90 (95% CI: [0.60, 1.35], P = 0.62) for those with no alternate rs4645 alleles (**Fig 3c**).

We suspected that linkage disequilibrium underlies the observed interaction, as the enhancing modifier rs4645 and *HFE* are both on chromosome 6 (**Fig 3d**). Using the 1000G Project, we found that rs4645 is in linkage disequilibrium with C282Y (R^2^ = 0.14, D’ = 0.98), but not with H63D (R^2^ = 0.66, D’ = 0.003). When we modeled the interactions of C282Y and rs2858993 (vQTL) jointly with rs2858993’s interaction with rs4645, the interaction between C282Y and rs2858993 was significant (β_int_ = 0.03 g/dL, 95% CI = [0.02, 0.05], P = 1.08×10^-4^) while the interaction between rs4645 and rs2858993 was not (β_int_ = -0.003 g/dL, 95% CI = [-0.014, 0.0086], P = 0.61). Therefore, linkage disequilibrium with C282Y likely explains rs4645’s significant interactions with vQTLs in *HFE* and rs1799945 (**Supplementary Fig 3**). It is known that C282Y causes more severe type 1 hereditary hemochromatosis than H63D,^23^ and these data suggest that differences in pathogenicity between hereditary hemochromatosis variants explain the observed cis epistasis between rs4645 and the *HFE* variants.

Second, we examined the interaction hits of rs17425819, a vQTL in *JAK2* for platelet count. rs17425819 had a significant variance effect (β_dispersion_ = 0.050 10^9^ cells/L, 95% CI = [0.013, 0.087], P = 7.65×10^-3^). rs35417585 had a significant negative interaction with rs17425819 (**Fig 4a**, β_int_ = -1.13 10^9^ cells/L, 95% CI = [-1.57, -0.69], P = 4.37×10^-7^). In contrast to the cis interaction observed between the *HFE* variants on chromosome 6, the interaction occurring between rs17425819 on chromosome 9 and rs35417585 on chromosome 14 exemplifies a trans interaction. The vQTL rs17425819 is in linkage disequilibrium with rs59384377 which tags the *JAK2* 46/1 haplotype (**Fig 4b**; D’ = 0.93, R^2^ = 0.86). The 46/1 haplotype predisposes individuals to somatic mutations in *JAK2* and myeloproliferative neoplasms such as polycythemia vera (PV) via the *JAK2* V617F somatic mutation.^24–26^ Unsurprisingly, given strong linkage disequilibrium between the vQTL and rs59384377, rs59384377 also had a significant negative interaction with rs35417585 (**Supplementary Fig 4**; P = 1.39×10^-6^). In 70,745 individuals in BioVU, rs35417585 and rs59384377 also had a significant negative interaction for platelet count (β_int_ = -0.49 10^9^ cells/L, 95% CI = [-0.82, -0.16], P = 0.001). The interaction did not replicate in 91,946 individuals in All of Us with available data (β_int_= 0.71 10^9^ cells/L, 95% CI = [-0.384, 1.80], P = 0.20).

**Fig. 4:**
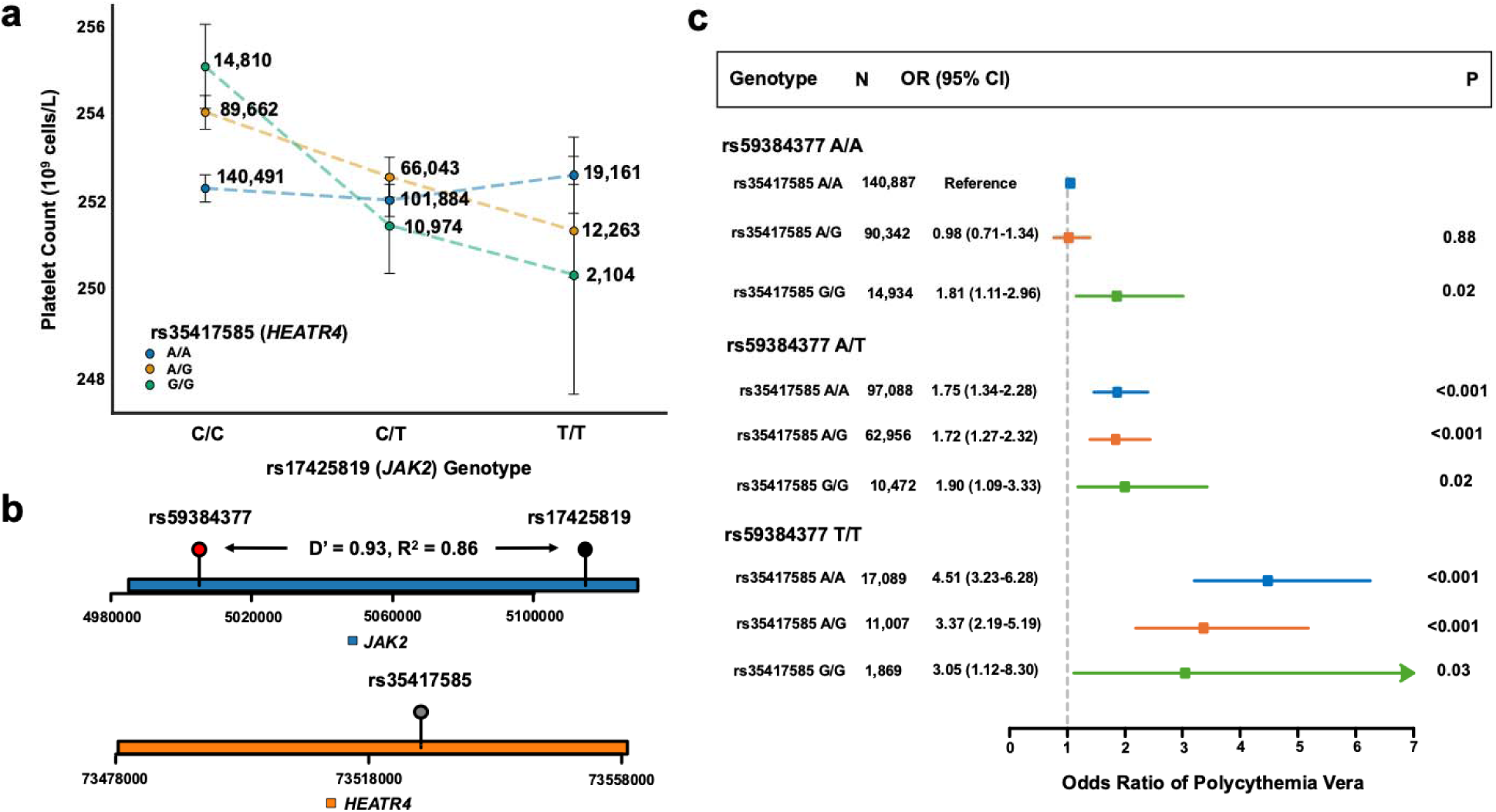
JAK2 and HEATR4 variants affect platelet count and polycythemia vera risk. a) Platelet count (10^9^ cells/L) stratified by genotypes at rs17425819 (*JAK2*) and rs35417585 (*HEATR4*). The x-axis shows rs17425819 genotypes (C/C, C/T, T/T), with separate lines for rs35417585 genotypes (A/A in blue, A/G in orange, G/G in green). The y-axis shows mean platelet count. Error bars represent 95% confidence intervals (1.96*SE). Numbers beside each point indicate sample sizes for each genotype combination. b) Genomic context of rs59384377 and rs17425819 in *JAK2*, and rs35417585 in *HEATR4*. The plot shows the relative positions of these variants within their respective genes. The x-axis indicates the base position on the chromosome. rs59384377 and rs17425819 are in linkage disequilibrium (D’ = 0.93, R^2^ = 0.86). c) Forest plot of odds ratios for polycythemia vera risk by rs59384377 (*JAK2*) and rs35417585 (*HEATR4*) genotype combinations. The x-axis shows the odds ratio. For each combination, the plot shows the number of individuals (N), odds ratio with 95% confidence interval (CI), and P-value (P). The rs59384377 A/A and rs35417585 A/A combination serves as the reference group.

We then sought to determine if rs35417585 also reduces risk of PV among people with the 46/1 haplotype in the UK Biobank. rs35417585 was not associated with PV alone (P=0.07), and the interaction between rs35417585 and rs59384377 was significant (β_int_= -0.21, 95% CI = [-0.43, - 6.0×10^-5^], P = 0.04), with rs35417585 associated with reduced risk of PV among those with the 46/1 haplotype (**Fig 4c**). Among those homozygous for the *JAK2* 46/1 haplotype, the odds of PV was 4.51 (95% CI = [3.23, 6.28], P < 0.001) for individuals homozygous for the rs35417585 reference allele. Odds decreased to 3.37 (95% CI = [2.19, 5.19], P < 0.001) and 3.05 (95% CI = [1.12, 8.30], P = 0.03) for rs35417585 heterozygotes and rs35417585 alternate allele homozygotes respectively. This interaction with PV is likely driven by *JAK2* V617F clonal hematopoiesis, defined as a copy-neutral loss of heterozygosity of the p-arm of chromosome 9 or a somatic single nucleotide variant causing *JAK2* V617F CHIP.^27–29^ Among 29,965 homozygotes for the *JAK2* 46/1 haplotype, each additional copy of the rs35417585 alternate allele reduced risk of JAK2 *V617F* clonal hematopoiesis by 1.25-fold (95% CI = [1.12, 1.75]).

Literature evidence suggests that alterations to fatty acid metabolism in PV and *JAK2* clonal hematopoiesis may contribute. rs35417585 is nearest to the *HEATR4* gene, but is also a splicing and expression quantitative trait locus of *ACOT1*, *ACOT2*, and *ACOT4*.^19^ The *ACOT* gene family encodes Acyl-CoA thioesterases, which hydrolyze coenzyme A esters into free fatty acids and coenzyme A. Prior work studying the metabolic differences of peripheral blood sera between 32 PV patients and 20 healthy controls suggests that derangements to fatty acid metabolism may provide a proliferative advantage in PV.^30^ Genetic variation affecting expression of the *ACOT* family may provide protection against PV in some individuals with the *JAK2* 46/1 haplotype.

Finally, we used the theoretical expectation of additive-by-additive effect size range in the GWAS SNP-SNP interaction model to determine if we had power to detect pairwise interactions in the UK Biobank, as Jabalameli et al performed in 23AndMe for height.^10^ We estimate that we have over 80% power to detect an interaction effect of magnitude approximately 0.05 times the standard deviation of the trait for minor allele frequencies about 10% and 80% power to detect an interaction effect of magnitude approximately 0.5 times the standard deviation of the trait for minor allele frequencies of 1% (**Supplementary Fig 4**). This suggests that the study’s large sample size is sufficient to ensure a high probability of detecting gene-by-gene interactions up to minor allele frequencies of 10%; indeed, all of the interactions discovered in this cohort were between variants with minor allele > 10%.^31^

This work has several limitations. First, the study uses data from predominantly individuals of European ancestry (i.e., UK Biobank) for the discovery of vQTLs and gene-gene interactions. The inclusion of multiple ancestries when using vQTLs to find gene-gene interactions can increase type 1 errors, as variants with differing allele frequencies by ancestry may be vQTLs without any underlying interaction.^32^ Second, we chose a conservative threshold of mAF > 10% to examine variants, as we are underpowered to detect interactions for less common variants. As more individuals are sequenced, the power to detect gene-gene interactions will increase, enabling us to study interactions between rarer variants.

In conclusion, our study demonstrates the utility of vQTLs in prioritizing gene-gene interactions for blood traits and disease risk in the UK Biobank. We identified significant interactions affecting hemoglobin levels, platelet count, and eosinophil and lymphocyte counts and disease risks for hereditary hemochromatosis, polycythemia vera, and celiac disease. While linkage disequilibrium with large main effect variants can explain apparent epistasis, such as for *HFE*, the interactions between *JAK2* and *HEATR4*, *CCL24* and *CCL26*, and *HLA-DQA1* and *HLA-DQB1* reveal novel modifiers of traits and disease risk. Future studies may apply vQTL-based approaches to a broader range of phenotypes and in more diverse populations. Ultimately, a more comprehensive understanding of gene-gene interactions may improve our ability to predict disease risk and advance personalized medicine.

## Methods

### UK Biobank Study Samples and Laboratory Data

The UK Biobank is a prospective epidemiological study, which includes genetics, health outcomes, and clinical laboratory measurements.^33^ We included individuals with phenotypes for the following laboratory measurements: platelet count (field ID: 30080), hemoglobin (field ID: 30010), eosinophil count (field ID: 30150), and lymphocyte count (field ID: 30120). Data was accessed and analyses were performed on UK Biobank’s Research Analysis Platform.

### UK Biobank Genetic Data

The directly genotyped variants were from release version 2, performed using the Applied Biosystems UK BiLEVE Axiom array (n = 49,950) or the Applied Biosystems UK Biobank Axiom array (n = 438,427). Imputed variants were inferred using the TOPMed reference panel from release 3. Both were aligned to GRCh37. The phased variants were from a dataset containing 200,031 samples with whole genome sequencing aligned to GRCh38 (cite phasing doc).

### UK Biobank Proteomic Data

The UK Biobank performed the Olink proteomics assay on 54,219 individuals. Further details of the Olink proteomics assay, data processing and quality control can be found in prior papers.^17^ Each protein level was inverse-rank normalized before analyses and association testing. All available individual-level plasma proteomics values were extracted for CCL24 and CCL26. Association testing adjusted for age, age^2^, sex, genetic ancestry, the first 5 genotyping principal components, and proteomic batch effects.

### Diagnoses

Polycythemia vera was defined by the ICD10 code D45. Cirrhosis was defined as ICD10 codes: K74.5, K74.3, K74.0, and K74.6.^34^ Celiac disease was defined by the ICD10 code K90.

### Variant Quantitative Trait Loci Testing with dglm

Variance quantitative trait loci (vQTLs) were identified using dglm v.1.8.6 in R version 4.4.0. The package dglm uses one generalized linear model (GLM) to fit the specified response and a second GLM to fit the deviance of the first model. vQTLs in each gene of interest (**Supplementary Table 4**) were identified if they had a significant dispersion p-value after Bonferroni correction (0.05/number of variants in the gene with minor allele frequency > 10%). The models were adjusted for age at blood draw, age at blood draw^2^, sex, genetic ancestry, and the first 5 genotyping principal components.

### Single Variant Association

All single variant associations were performed using an executable in the DNAnexus platform. The executable implements Regenie (v3.3), a linear mixed model, which incorporates both fixed and random effects to account for population stratification and relatedness.^14^ A random selection of 500,000 variants for step one with a minor allele count >5,000 were included for step one.

Variants with a minor allele frequency (mAF) < 0.001 and a genotyping rate < 0.1 were excluded for the second step. Samples that did not report assigned male or female at birth were also excluded. All laboratory traits were rank-based inverse-normal transformed and used at the dependent variable. Age at blood draw, age at blood draw^2^, genetic sex, and the first ten ancestry principal components were included as covariates.

### Scale Test: Genome-Wide Associations with Variance

To perform the Scale test, we rank inverse normal transformed the blood traits using scipy v1.14.1. Then, we performed two single variant association models with Regenie v3.3. The first phenotype was for the rank-inverse-normal-transformed trait, which identified variants associated with a mean effect. The second phenotype was the square of the residual of the rank-inverse-normal-transformed trait after adjusting for age at blood draw, age at blood draw^2^, sex, genetic ancestry, and the first 5 genotyping principal components; this phenotype identified traits associated with a variance effect. A variant was identified as a vQTL if it had a genome-wide significant association with the variance of a trait (P < 5×10^-8^) without a significant mean effect after multiple-hypothesis correction (P > 5×10^-8^). vQTLs were then extracted and tested in an interaction model.

### Regenie Interaction Modeling

All interaction modeling was performed using the executable, variant lists, and covariates implemented in the single variant association. In the second step of the process, REGENIE also conducted an interaction scan between the test variants and interaction variant, employing an additive model.

### Variant Extraction

Genotype calls were extracted from multi-sample BGENs using Plink (v2.00a3.1LM), loaded in an executable available in the DNAnexus platform. Variant calls were extracted from the multi-sample phased vcfs using bcftools (v.1.15.1). Variants with a minor allele frequency >10% were extracted from genes of interest (**Supplementary Table 4**) for dglm modeling. Variants that were hits from the interaction modeling were extracted for plotting and sensitivity analyses.

### JAK2 Clonal Hematopoiesis Detection

Individuals with *JAK2* clonal hematopoiesis in the UK Biobank were identified based on (1) somatic mutations detected from whole exome sequencing data in JAK2 V617F, as described in Vlasschaert et al or (2) the mosaic chromosomal alteration of copy-neutral loss of heterozygosity on the p-arm of chromosome 9 using genotyping array data as described in Loh et al.^27,28^

### BioVU Validation Cohort

BioVU is Vanderbilt’s biorepository of DNA extracted from discarded blood collected during routine clinical testing. The genomic data is linked to de-identified medical records in the Synthetic Derivative. 70,745 people in Vanderbilt’s BioVU cohort were genotyped on the Infinium Multi-Ethnic Genotyping Array (MEGAchip).^35^ We included individuals with the following laboratory measurements: platelet count (OMOP Concept ID: 3024929), hemoglobin (OMOP Concept ID: 3000963), eosinophil count (OMOP Concept ID: 3028615), and lymphocyte count (OMOP Concept ID 3004327). We used the median of all of an individuals’ laboratory values if an individual had multiple values. We excluded laboratory values greater than 3-fold above the upper limit of normal, since BioVU is a hospital-based cohort (e.g., individuals may have infections causing elevated lymphocyte counts not representative of their normal lymphocyte count). Data was accessed and analyses were performed on BioVU’s Terra.bio platform.

### All of Us Validation Cohort

The All of Us Research Program is a biobank that collects a wide range of data from participants, including detailed health information, biological samples, and genomic data. Sequencing reads were aligned to the GRCh38 reference genome. Informed consent for all participants was conducted either in person or through an eConsent platform approved by the Institutional Review Board (IRB) of the All of Us Research program. We identified 138,938 individuals with whole-genome sequencing data and available laboratory values (same laboratory measurements and OMOP Concept IDs as in BioVU). Data was accessed and analyses were performed on the NIH All of Us Researcher Workbench cloud platform.

### Linkage Disequilibrium using LDLink

Variant R^2^ and D’ values were queried using LDLink’s LDpair tool using all available populations (https://ldlink.nih.gov/?tab=ldpair).

### Statistical and Plotting Methods

Lollipop plots were generated using trackViewer version 1.40.0 in R version 4.4.0.

### Power Analysis

Power calculations were based on the non-central chi-squared distribution. We considered a range of minor allele frequencies (MAFs) and interaction effect sizes, in units of standard deviations, to explore how these parameters influence power. The effect sizes of the two individual SNPs were set to 0.01. For each combination of MAFs and interaction effect size, we calculated the expected mean and variance of the trait, SNP, and interaction term under the null hypothesis of no interaction. Then, using these parameters, we estimated the non-centrality parameter and calculated power as the probability of rejecting the null hypothesis given the non-centrality parameter. We generated power curves for a range of interaction effect sizes as a function of the MAFs of the two SNPs. These analyses were performed assuming a sample size of 400,000 (similar to the UK Biobank), a significance level of 5×10^-8^, and a normally distributed trait with a mean of 0 and a standard deviation of 1.

## Data Availability

Genetic data is available for the UK Biobank at https://ukbiobank.dnanexus.com, for NIH AllofUs at https://workbench.researchallofus.org, and BioVU data on the Synthetic Derivative on app.terra.bio. Supplementary tables are available here: https://docs.google.com/spreadsheets/d/1SJelCr6tP6devAAgFhpBWeq1XBVC8bbEbZzkk2Ql-2g/edit?usp=sharing.

## Code Availability

Code is available at https://github.com/bicklab/gxg-interaction-modeling. Information about regenie is available at https://rgcgithub.github.io/regenie.

