## Supplementary Materials for "Variance quantitative trait loci reveal gene-gene interactions which alter blood traits"

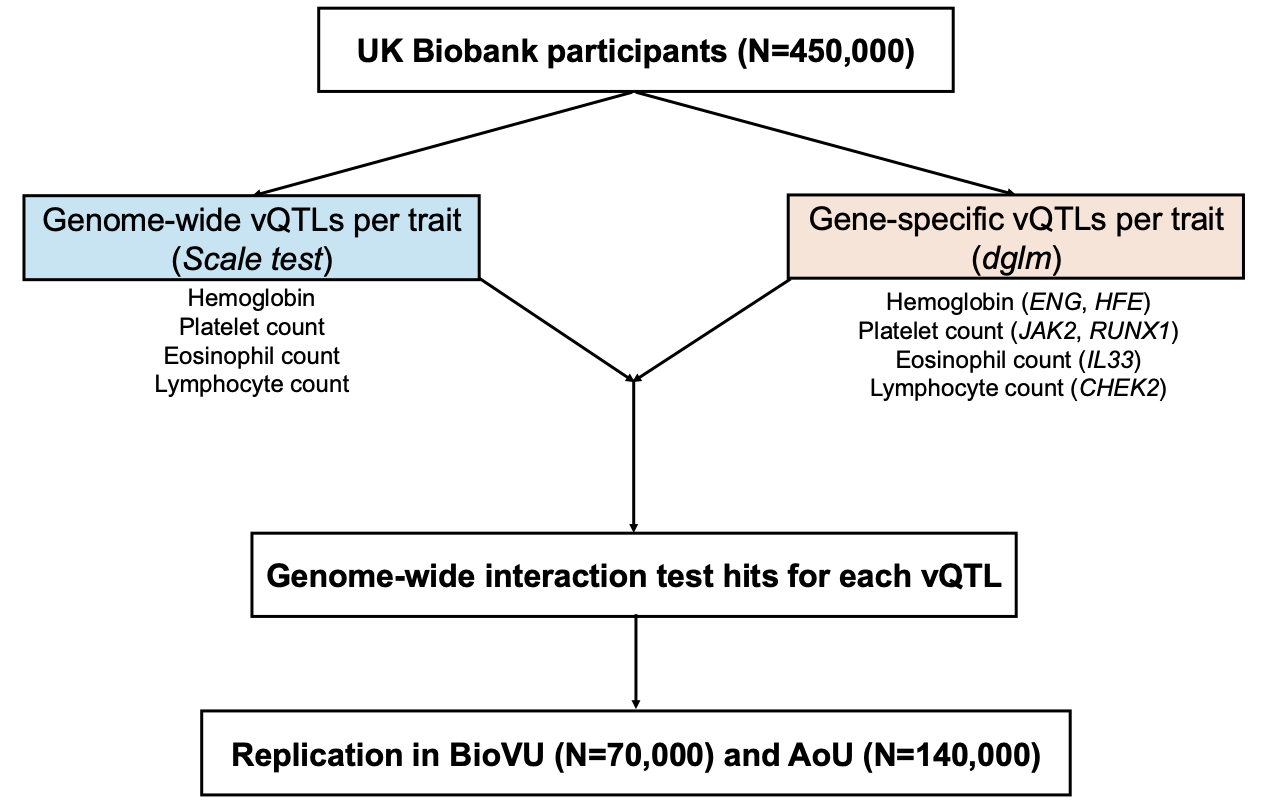

**Supplementary Fig 1: Study design.** In the UK Biobank (UKB), there are approximately 450,000 individuals with genotyping array data and index laboratory values for platelet count (field ID: 30080), hemoglobin (field ID: 30010), eosinophil count (field ID: 30150), and lymphocyte count (field ID: 30120). First, we performed the Scale test which identifies genome-wide putative variance quantitative trait loci (vQTLs) which are not statistically significant in a genome-wide association study (GWAS) of a trait but is significant in a GWAS of the square of the residuals. Second, we sought to identify vQTLs in disease-relevant genes in order to improve our power to detect vQTLs. We modeled the mean and variance with double-generalized linear models (dglm) and identified gene-specific vQTLs per trait. For each vQTL discovered through either method, we performed genome-wide interaction testing. Then, for each significant interaction, we extracted the pair of variants in approximately 70,000 people in Vanderbilt’s BioVU cohort and 140,000 people in NIH’s All of Us (AoU) cohort.

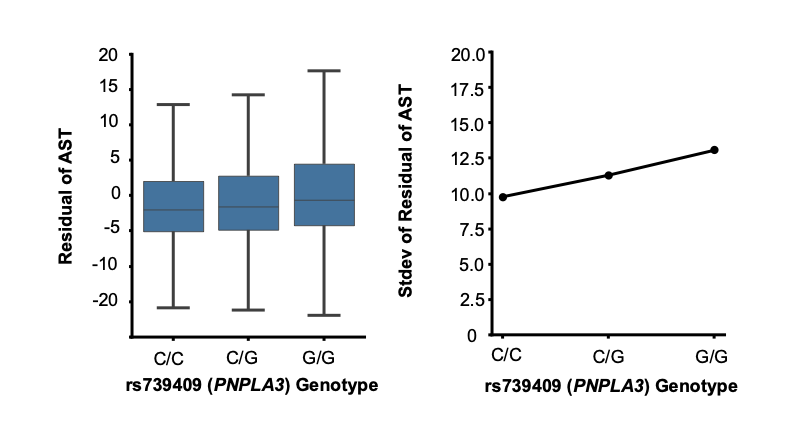

**Supplementary Fig 2: rs739409 in *PNPLA3* is a variance quantitative trait locus.** Left: Residual AST levels stratified by rs739409 genotype (C/C, C/G, G/G). N=299,550 C/C individuals, N=164,321 C/G individuals, and N=23,200 G/G individuals. Each dot represents an individual data point, with the box plot showing median, interquartile range, and whiskers extending to 1.5 times the interquartile range. The y-axis shows residual AST values after adjusting for age, age2, sex, and genetic principal components 1 through 5. Right: Standard deviation of residual AST levels for each rs739409 genotype group.

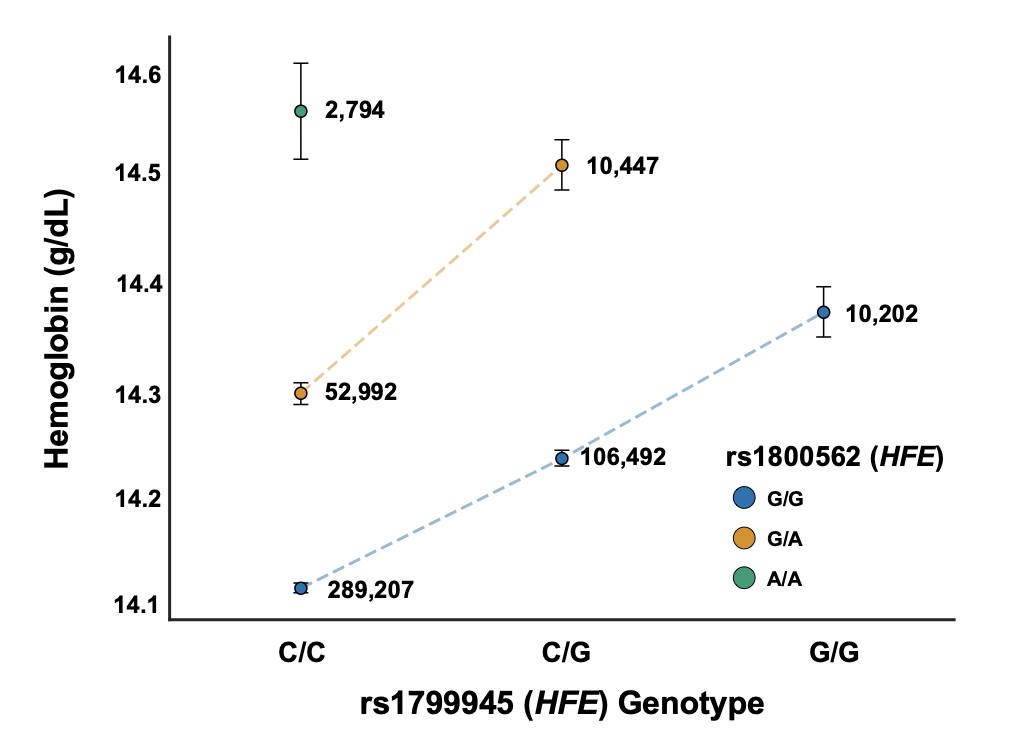

**Supplementary Fig 3: Interaction between HFE variants rs1799945 and rs1800562 affects hemoglobin levels.** The figure shows hemoglobin levels (g/dL) stratified by genotypes at rs1799945 and rs1800562, both located in the *HFE* gene. The x-axis represents rs1799945 genotypes (C/C, C/G, G/G), while different colors indicate rs1800562 genotypes (G/G in blue, G/A in orange, A/A in green). The y-axis shows mean hemoglobin levels. Each data point represents a specific genotype combination, with error bars indicating one standard deviation (1.96*SE). The number next to each point shows the sample size for that genotype combination. Dashed lines connect points of the same rs1800562 genotype across different rs1799945 genotypes.

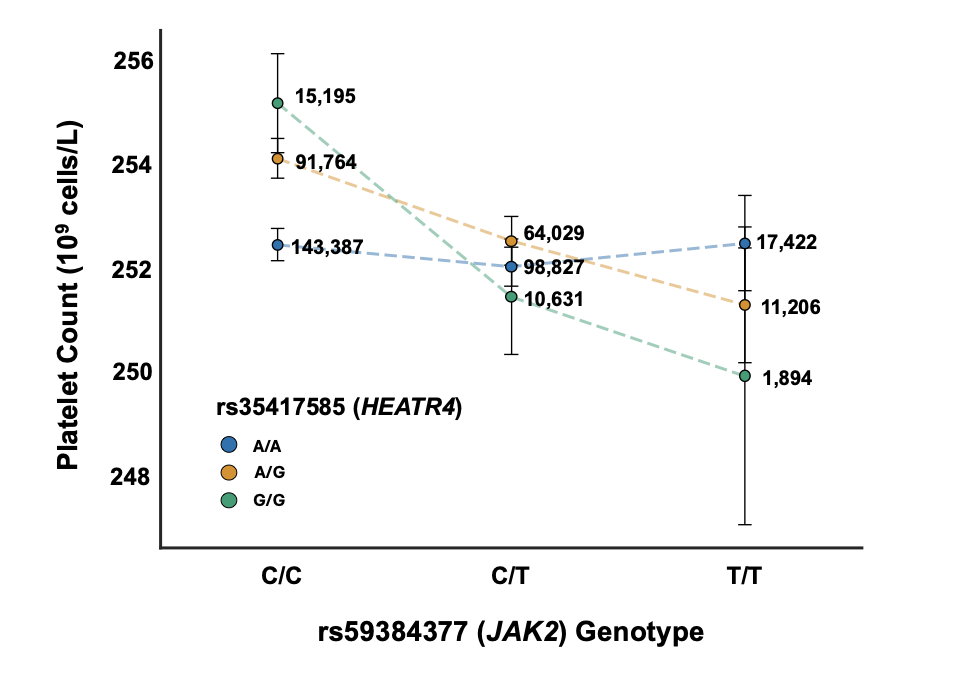

**Supplementary Fig 4: Interaction between JAK2 variant rs59384377 and HEATR4 variant rs35417585 affects platelet count.** The figure shows platelet count (10^9^ cells/L) stratified by genotypes at rs59384377 (*JAK2*) and rs35417585 (*HEATR4*). The x-axis represents rs59384377 genotypes (C/C, C/T, T/T), while different colors indicate rs35417585 genotypes (A/A in blue, A/G in orange, G/G in green). The y-axis shows mean platelet count. Each data point represents a specific genotype combination, with error bars indicating one standard deviation (1.96*SE). The number next to each point shows the sample size for that genotype combination. Dashed lines connect points of the same rs35417585 genotype across different rs59384377 genotypes.

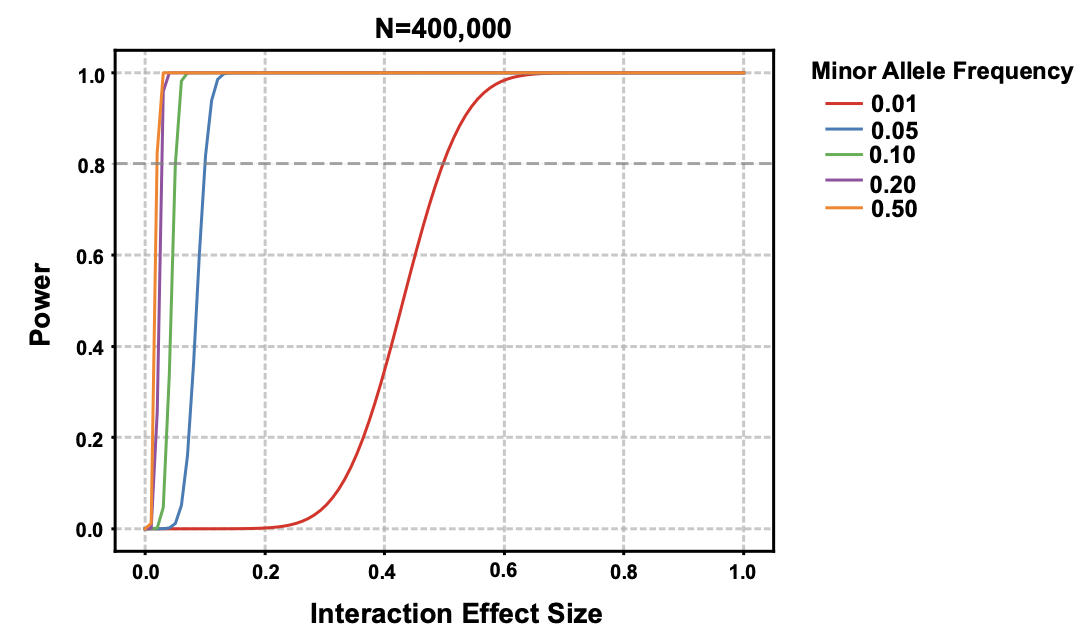

**Supplementary Fig 5: Power analysis for detecting genetic interactions at varying effect sizes and minor allele frequencies.** This figure illustrates the statistical power to detect genetic interactions in a sample size of 400,000 individuals across different interaction effect sizes and minor allele frequencies (MAF). The x-axis represents the interaction effect size, ranging from 0 to 1. The y-axis shows the statistical power, also ranging from 0 to 1. Each line on the plot represents a different minor allele frequency, color-coded as follows: Red: MAF = 0.01, Blue: MAF = 0.05, Green: MAF = 0.10, Purple: MAF = 0.20, and Orange: MAF = 0.50.
